# Effect of a continuous trauma quality improvement programme on mortality in urban India: a non-randomised controlled trial

**DOI:** 10.1101/2024.05.27.24307748

**Authors:** Johanna Berg, Siddarth David, Girish D. Bakhshi, Debojit Basak, Shamita Chatterjee, Kapil Dev Soni, Ulf Ekelund, Li Felländer-Tsai, Manjul Joshipura, Tamal Khan, Monty Khajanchi, Mohan L N, Anurag Mishra, Max Petzold, Sendhil Rajan, Nobhojit Roy, Rajdeep Singh, Martin Gerdin Wärnberg

## Abstract

**Background:** Trauma is the leading cause of quality-related mortality in low- and middle-income countries, with an estimated two million preventable deaths each year. Although trauma quality improvement programmes have been utilised in high-income countries for more than three decades, there is no high-level evidence of their effect on patient outcomes. We aimed to assess whether implementing a continuous trauma quality improvement programme using audit filters improves mortality among adult trauma patients in urban India.

**Methods:** We conducted a prospective controlled interrupted time-series study across four tertiary hospitals in urban India between 2017 and 2022. Two hospitals implemented a trauma quality improvement programme after a one-year observation phase (intervention arm); two continued standard care (control arm). Time-series analysis was done using monthly aggregated data with generalised additive models. A difference-in-differences approach was used for secondary analysis. The primary outcome was all-cause in-hospital mortality; the secondary outcome was 30-day mortality.

**Findings:** In total, 10 143 adult trauma patients were included (median age 35 years; 83% men). In-hospital mortality decreased by 11.2% (95% CI –16.0 to –5.5) in the intervention arm, with no evidence of change in the control arm (–0.5%; 95% CI –4.0 to 5.4). Secondary difference-in-differences analysis showed consistent reductions in in-hospital mortality (–12%; 95% CI –16 to –9) and 30-day mortality (–15%; 95% CI –19 to –11). No seasonal or autocorrelation effects were observed. External factors, including the opening of a dedicated trauma centre at one site and the COVID-19 pandemic, may have influenced the results.

**Interpretation:** Trauma quality improvement programmes may improve mortality, particularly in settings with a high number of preventable deaths, and can contribute to reducing the global burden of quality-related mortality. Further research is needed to confirm these results, clarify the mechanisms through which such programmes mediate their effect, and determine how they can be sustained over time.

**Funding:** This work was funded by the Swedish Research Council (2016-02041).

**Trial registration:** Trauma Audit Filter Trial, ClinicalTrials.gov ID NCT03235388, https://clinicaltrials.gov/study/NCT03235388

**Research in context Evidence before this study:** We searched MEDLINE, Embase, Web of Science, Cochrane CENTRAL, CINAHL, and PubMed, plus ClinicalTrials.gov (English only records), using combinations of (“trauma” OR “injury”) AND (“audit filter” OR “quality indicator”). We found no randomised or quasi-experimental studies assessing the effect of trauma quality improvement programmes on mortality. Two prospective before–after studies were identified: one from Thailand (2001) reporting a reduction in preventable deaths and improved care processes, and one from Germany (2002) showing process improvements with a non-significant reduction in mortality. These studies were included in the review that informed the 2009 WHO Guidelines for Trauma Quality Improvement Programmes.

**Added value of this study:** This is the first quasi-experimental study to assess whether implementing a trauma quality improvement programme using audit filters reduces mortality. The programme was implemented in two tertiary hospitals in urban India, with two hospitals serving as controls. Implementation was associated with significant reductions in in-hospital and 30-day mortality. This study provides the first high-level evidence that a data driven continuous trauma quality improvement program, as outlined in the WHO guidelines, may improve mortality in adult trauma patients.

**Implications of all the available evidence:** Trauma quality improvement programmes may reduce mortality, particularly in settings with high numbers of preventable deaths. The available evidence supports the implementation and scale-up of structured, continuous trauma quality improvement programmes to reduce quality related trauma mortality in LMICs. Further research is needed to confirm these findings in other settings, and to identify the mechanisms through which such programmes achieve and sustain their effects.

## Introduction

Improving quality of care remains essential, as poor quality is a greater cause of preventable deaths worldwide than lack of access to services.^1^ Quality improvement initiatives are frequently proposed to enhance patient safety and improve care,^2^ but the evidence for their effectiveness in improving patient outcomes remains limited. Studies report conflicting results and often lack theoretical foundations that explain their mechanism of action.^2–4^ Trauma results in more than four million deaths annually and accounts for the largest number of quality related deaths in low- and middle-income countries (LMICs).^1,5^ More than half of trauma deaths are caused by poor quality among those who reach care.^1^ Over two million lives could be saved each year if mortality rates in low- and middle-income countries (LMICs) were the same as those in high-income countries (HICs).^6^

Strengthening the quality of trauma care is thus critical to reduce preventable trauma deaths and global inequities in survival.^1^ Attempting to address these inequities, WHO released guidelines for trauma quality improvement programmes in 2009, though widely applied in HIC, there is no high-level evidence that trauma quality improvement programs improves outcomes, and uptake and dissemination have been slow and uneven across LMICs.^7,8^

These programmes include identifying and analysing departures from care standards, evaluating opportunities for improvement, and implementing corrective action plans to improve patient care.^7,9,10^ Identifying cases with deviations from care is traditionally done using audit filters. They are predefined statements that represent consensus based ideal care standards, for example, “Patients with a Glasgow Coma Scale (GCS) score of less than 8 should receive a definitive airway”.^7,11^

The American College of Surgeons released the first trauma audit filters as part of the guidelines on trauma care in 1987.^12^ Since then, the use of audit filters and trauma quality improvement programmes has been widely applied in HICs, and in 2009 they were recommended by the WHO as one way to mitigate the inequalities of trauma care globally — despite there being no high-level evidence for their effectiveness in improving patient outcomes.^13^ Our aim was therefore to assess whether implementing a trauma quality improvement programme using audit filters improved patient outcomes.

## Methods

### Study design and setting

We conducted a controlled interrupted time series study across four hospitals in urban India (ClinicalTrials.gov identifier NCT03235388^14^). All tertiary care hospitals have approximately 1500 beds and in-house clinical specialities to care for trauma patients. None of the hospitals used any structured trauma quality improvement process prior to this study, and no electronic health records. The study had three phases: an observation phase lasting 14 months in all four hospitals to establish baseline outcomes, an implementation phase lasting six months, during which two hospitals were randomized to implement a trauma quality improvement programme with audit filters, and an intervention phase lasting 41 months. The intervention phase was extended by 18 months due to the COVID-19 pandemic. We performed an interim analysis 15 months after the initiation of the intervention to asses the quality of the data and identify unexpected changes in outcomes. We followed the TREND checklist for reporting of non-randomised clinical trials.^15^

### Intervention and control

For the intervention centres, a multidisciplinary review group was established at each centre, including representatives from all departments involved in trauma care: emergency/casualty officers, general surgery, anaesthesia, burns and plastics, orthopaedics, radiology, and neurosurgery. The local principal investigator at each centre recruited senior healthcare staff from each department. Within this group, an anonymous online Delphi process was conducted to identify which audit filters would be most useful to implement at each hospital.^16^ Further details on the Delphi process, including participant composition and procedures, are available in the cited publication. A list of implemented audit filters is included as supplementary material.

A project officer prospectively included patients and identified specific audit filter violations through direct observation and data collection. Structured data were collected on violations, process outcomes, treatments, interventions, and patient outcomes during the hospital stay. These cases were compiled into automated structured reports every 4–6 weeks, which served as the basis for discussions at the multidisciplinary board review meetings. During each meeting, cases were reviewed and improvement actions identified. Progress on these actions was followed up in subsequent meetings. During the implementation phase, members of the core research team with experience in facilitating review discussions attended each meeting to guide and support the discussions, after this, only the local review board attended.

In the control centres, no intervention was implemented. They continued treating trauma patients using standard care, without a quality improvement process. The centre-specific data collected for this study were available to both intervention and control centres but were not used in any structured way in the control centres to inform or direct care. Weekly calls were held between the core research team and project officers at each hospital to support data collection and address operational issues. In intervention centres, multidisciplinary review meetings were conducted independently by the local teams, with no involvement from the research team beyond facilitating the underlying reports based on collected data.

### Participants

#### Inclusion criteria and enrollment

We included adults aged 18 years and older admitted for in-hospital care with a history of trauma, defined by International Classification of Diseases, tenth revision (ICD-10) chapter 20, codes V01-Y36 for external causes of morbidity and mortality, as the reason for admission. Project officers were trained to record vital signs with standard equipment for patients across rotating shifts (day, evening, and night). Each shift comprised six hours for enrolling new patients in the emergency department and two hours for follow-up of previously included patients.

#### Retrospective inclusion

Due to lower than expected prospective inclusion rates, particularly during the COVID-19 pandemic, we retrospectively included patients to achieve the required sample size for the primary statistical analysis. Using hospital records, we randomly selected trauma patients admitted each month. As informed consent could not be obtained, only basic, reliably recorded data were collected: date of admission, age, sex, mechanism of injury, and in-hospital mortality. Data were extracted from ward books and paper charts, as no electronic health record was available.

#### Data collection and quality control

Initial data collection was performed using paper forms, and the data were then periodically entered into a digital data collection tool and uploaded to secure servers. Patient identifiers were not uploaded, and identification was only possible through the original paper records, which were stored at each hospital in compliance with local regulations. Basic data validation was performed at the time of entry, and double entry of all variables was performed after all the data had been collected to minimize the risk of transfer errors. Quality control visits were conducted each quarter with project officers from different hospitals evaluating the data collection processes based on predefined criteria.

### Outcomes and covariates

Our primary outcome was all-cause in-hospital mortality. Information on 30-day and 90-day mortality was collected from medical records if the patient died during the hospital stay, otherwise, it was obtained through telephone follow-up. 30-day mortality, originally designated as the primary outcome, was analysed as a secondary outcome due to lower-than-expected prospective inclusion rates. We also collected data on additional secondary outcomes, including length of stay and intensive care unit (ICU) admissions. Data on several covariates, such as age, sex, vital signs, injury severity, and diagnostic procedures, were collected to describe and compare cohorts and, if needed, to adjust for case-mix and differences between hospitals.

### Study execution

One principal investigator and one co-principal investigator were recruited for each participating hospital. These were all experienced trauma clinicians with extensive local knowledge and research interest.

#### Observation phase (Month 1-14)

The participating hospitals started data collection at the same time to establish baseline outcomes. Weekly meetings were held with all project officers and the core research team to identify and address any issues related to data collection.

#### Implementation phase (Month 15-20)

The participating hospitals were paired so that the two hospitals with the highest and lowest volumes formed one pair, and the two remaining hospitals formed the second pair. One pair of hospitals was then randomly selected to implement the trauma quality improvement programme, becoming intervention hospitals, while the other two were control hospitals. The process of data collection remained the same at all sites.

At the intervention hospitals, a two-day session on the background and rationale of trauma audit filters was held by representatives from the core research team. This session was attended by the multidiciplinary team selected for the local review board as previously described. During the implementation phase, participants from the core research team attended audit filter review meetings to facilitate the discussion and formulate corrective strategies accordingly.

#### Intervention phase (Months 21-42)

The intervention hospitals continued the data collection both for base data and for audit filter deviations by two different project officers. The trauma audit review board continuously analysed deviations, implementing corrective strategies. The meetings were held without participation from the core research team. Three months after the intervention phase started, the largest intervention hospital opened a dedicated trauma centre. The control hospitals continued with base data collection.

### Statistical analysis

We collected both prospective and retrospective data, and the prospective cohort represents a random subset of the full study population. Some pre-defined analyses were therefore feasible only for the prospective cohort. Our primary analysis used a segmented interrupted time series model with a generalized additive model (GAM) to estimate the effect of the intervention on in-hospital mortality. To assess robustness, we conducted two secondary analyses: a difference-in-differences analysis using individual-level data and an unadjusted pre–post analysis.

### Primary analysis

For our primary analysis, we applied a segmented Generalized Additive Model (GAM) to estimate the effect of the intervention on mortality.^17^ Data were aggregated by study month across intervention and control arms, with each observation representing the average outcome for all patients enrolled during that month. The intervention was modelled as a step change, represented by a binary indicator. Time was included as a smooth term to flexibly capture potential non-linear trends. Calendar month was entered as a cyclic cubic spline with 12 knots to adjust for seasonality. The model used a beta regression distribution with a logit link, suitable for modelling bounded continuous outcomes between 0 and 1. Estimation was performed using restricted maximum likelihood (REML). Autocorrelation was assessed using the Box-Ljung test (lag = 12, degrees of freedom = 0), applied separately to response residuals from the intervention and control arms.

GAM model specification:

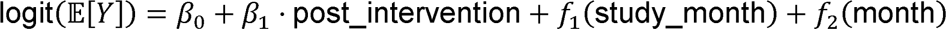

Here, *β_0_* is the intercept, and *β_1_* estimates the step change in mortality associated with the intervention. The smooth function *f_1_* (study_month) captures long-term, non-linear time trends across the study period, and the cyclic smooth *f_2_* (month) adjusts for recurrent seasonal variation. We developed a counterfactual model to compare predicted outcomes under scenarios without the intervention.

### Sample size considerations

The sample size requirements for interrupted time series analyses depend on several factors, including model complexity, data variance and the spread of observations. For our primary analysis, we chose to adhere to published guidelines stating that at least twelve observations are needed during the observation phase and twelve observations are needed during the intervention phase and that each observation should be an aggregate of at least 100 patients.^18^ Our use of a large GAM introduced extra complexity, and we estimated that 100 patients/arm/month and an extended intervention phase were needed to detect the potential impact of the intervention. This sample size allowed us to detect a reduction in mortality from 20% to 15% (power 0.8, alpha 0.05) using a pre-post design.

### Secondary and subgroup analysis

For the secondary analysis, we conducted a difference-in-differences (DiD), using linear regression on individual-level data for prospectively included patients. We adjusted for confounders selected based on clinical relevance and observed baseline differences between arms. To assess the parallel trends assumption, we plotted pre-intervention trends in in-hospital and 30-day mortality stratified by study arm, and visually inspected the trajectories. In addition, we tested for interaction between time (study month) and group (intervention vs control) using pre-intervention prospective data. All analyses were conducted in R^19^, using the _mgcv_ package for GAM modelling.

We conducted two pre-specified subgroup analyses for major trauma and potentially salvageable trauma patients. We defined a major trauma patient as any patient who was in the hospital for more than three days with an Injury Severity Score (ISS) >15 or who died within three days of arrival, and a potentially salvageable trauma patient as any patient with 15<ISS<24. Due to limited statistical power to conduct stratified interrupted time series analyses, these subgroup analyses were conducted using the secondary difference-in-differences approach. To account for multiple comparisons, we applied the Holm correction method.^20^

### Sensitivity analyses

We conducted two pre-specified sensitivity analyses: an unadjusted pre-post analysis using a two-sample Z-test for proportions, and an interrupted time series analysis excluding the implementation phase. Three post hoc analyses were also conducted: excluding the period after a trauma centre opened at one intervention hospital, excluding the COVID-19 phase, and performing unadjusted and adjusted interrupted time series regression in the prospectively included cohort in the intervention arm to evaluate whether observed effects on mortality were confounded by changes in case mix over time.

### Missing data and data processing

We added an analysis of missing data to the analysis plan to assess whether complete case analysis would be appropriate. We examined associations between missingness and key variables using the R package finalfit^21^, which applies chi-squared tests for categorical variables and univariate logistic regression for continuous variables, with missingness as the outcome. If no significant associations were found, indicating data were Missing Completely at Random (MCAR), we planned to proceed with complete case analysis. If significant associations were found, suggesting data were Missing at Random (MAR), we planned to use multiple imputation by chained equations (MICE), provided that the proportion of missing data was >5% and that the available data supported stable estimation of the imputation model. If data were deemed likely to be Missing Not at Random (MNAR), complete case analysis would be applied with caution, and sensitivity analyses would be considered. The Abbreviated Injury Scale (AIS) and Injury Severity Score (ISS) were calculated from ICD-10 codes using the R package icdpicr^22^.

### External events during the study period

During the study period, two external events occurred that may have influenced patient outcomes. First, three months after the intervention phase began, the largest intervention hospital opened a dedicated trauma centre. Although all surgical and radiology services for trauma were previously available, they were distributed across separate buildings and wards, with no dedicated units for trauma patients. With the opening of the trauma centre, these services were consolidated into a single 180-bed facility with in-house CT, operating theatres, and both ICU and general wards dedicated to trauma care. Nurses also received trauma-specific training prior to the opening.

Second, the COVID-19 pandemic vastly disrupted hospital operations at all sites. Prospective patient inclusion was halted between April and December 2020, and review meetings were paused until mid-2021. The second intervention hospital was converted into a COVID-19 facility for much of 2020 and 2021, effectively eliminating trauma admissions during this period. In addition to straining hospital capacity, the pandemic reduced population mobility and led to an overall decline in trauma incidence.

### Deviations from study protocol

The intended primary outcome was 30-day mortality, but this became unfeasible due to low prospective inclusion. We therefore defined in-hospital mortality as the primary outcome, for which the study was adequately powered. 30-day mortality was analyzed as a secondary outcome. We also added two non-prespecified analyses: an analysis of missing data and a difference-in-differences analysis. These were introduced because we lacked sufficient power for a time-series analysis of 30-day mortality and observed an unexpected baseline difference in mortality between study arms. The difference-in-differences approach allowed formal comparison between arms with adjustment for confounders. Not all protocol outcomes are reported here; notably, quality of life results will be published separately.

### Ethical considerations

The need for informed consent to participate in the intervention was not deemed applicable, and we were granted waivers of informed consent for recording vital signs, demographic parameters and in-hospital outcomes. The project officers obtained written consent for telephone follow-up. Ethical approval was granted by the Swedish Ethical Review Authority (approved 2017-06-07 2017/930-31/2), as well as by all local ethical review boards at each participating hospital (Maulana Azad Medical College (MAMC) - approved 2017-07-19 F.1/IEC/MAMC/(57/02/2017/No 113. SSKM/IPGME&R, Kolkata – approved 2017-08-21, IPGME&R/IEC/2017/396. JJ Hospital, Mumbai – approved 2017-08-22, No. IEC/Pharm/CT/111/A/2017. St. Johns, Bangalore – approved 2017-08-24, 160/2017).

### Data sharing

The code for this publication and an anonymised dataset from the clinical trial are available on Zenodo and GitHub under a Creative Commons Attribution 4.0 International License (CC BY 4.0)^23^.

## Results

We included 10143 patients between October 2017 and October 2022. Of these patients, 4126 were prospectively included (Figure 1). We included an average of 166 (SD 61) patients per month, 45 out of 61 months did not reach the target number of inclusions, due to low inclusion rates and lower number of patients during COVID-19.

**Figure 1:**
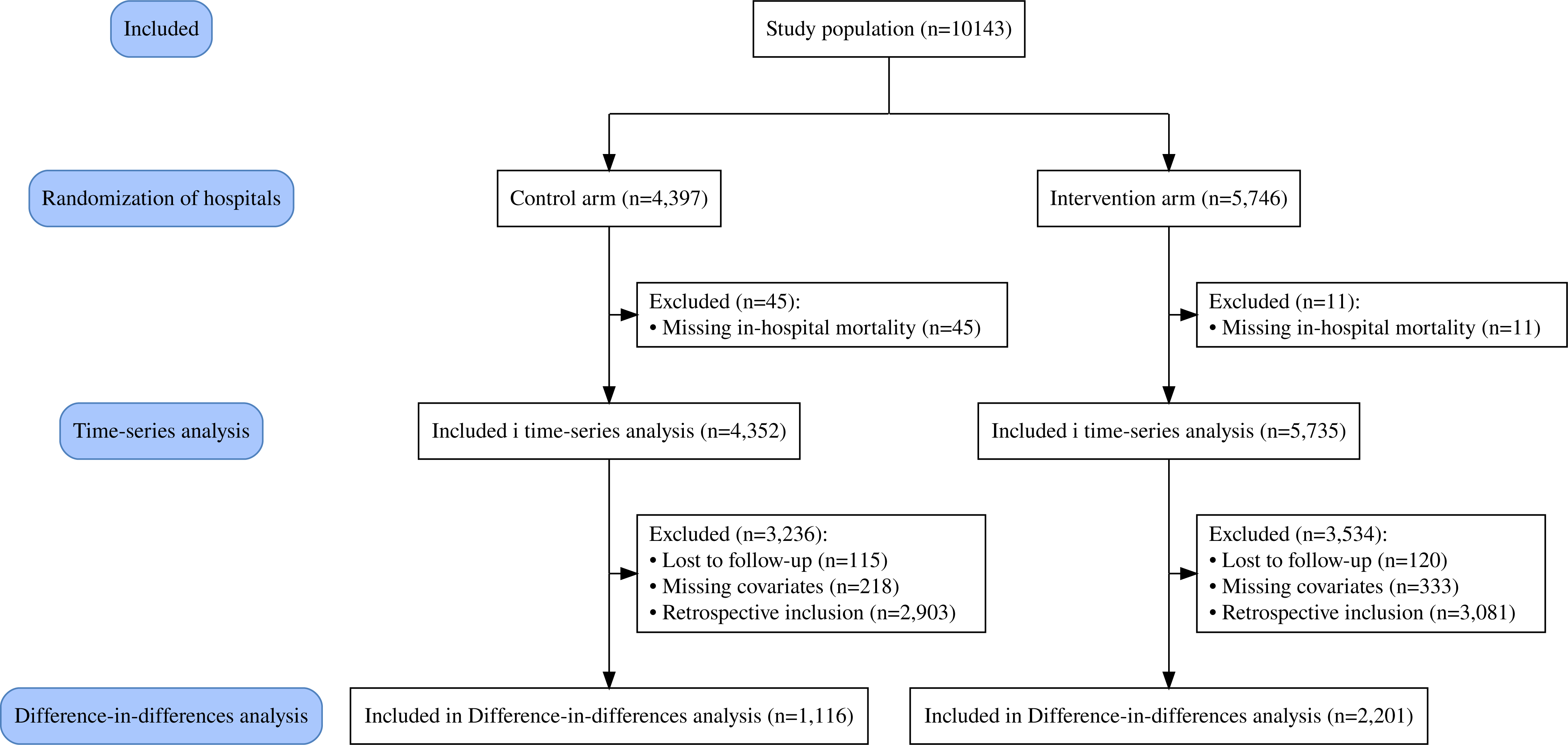
Flow chart.

Patient characteristics and missing data counts are presented in Table 1 (all patients) and Table 2 (prospectively included patients). For the primary analysis, missing data were minimal for in-hospital mortality (0.6%) and absent for study month. No significant associations were found between missingness and examined variables, except for one control arm month, which was deemed unlikely to affect results. In the secondary analysis using the adjusted DiD model, missingness was associated with GCS, ISS, and shock for 30-day mortality. The proportion of missing data was low for GCS (1.6%) and shock (2.2%), but higher for ISS (12.5%). Given the exploratory nature of the secondary analysis and the challenge of correctly imputing ISS, all statistical models used complete case analysis.

**Table 1:**
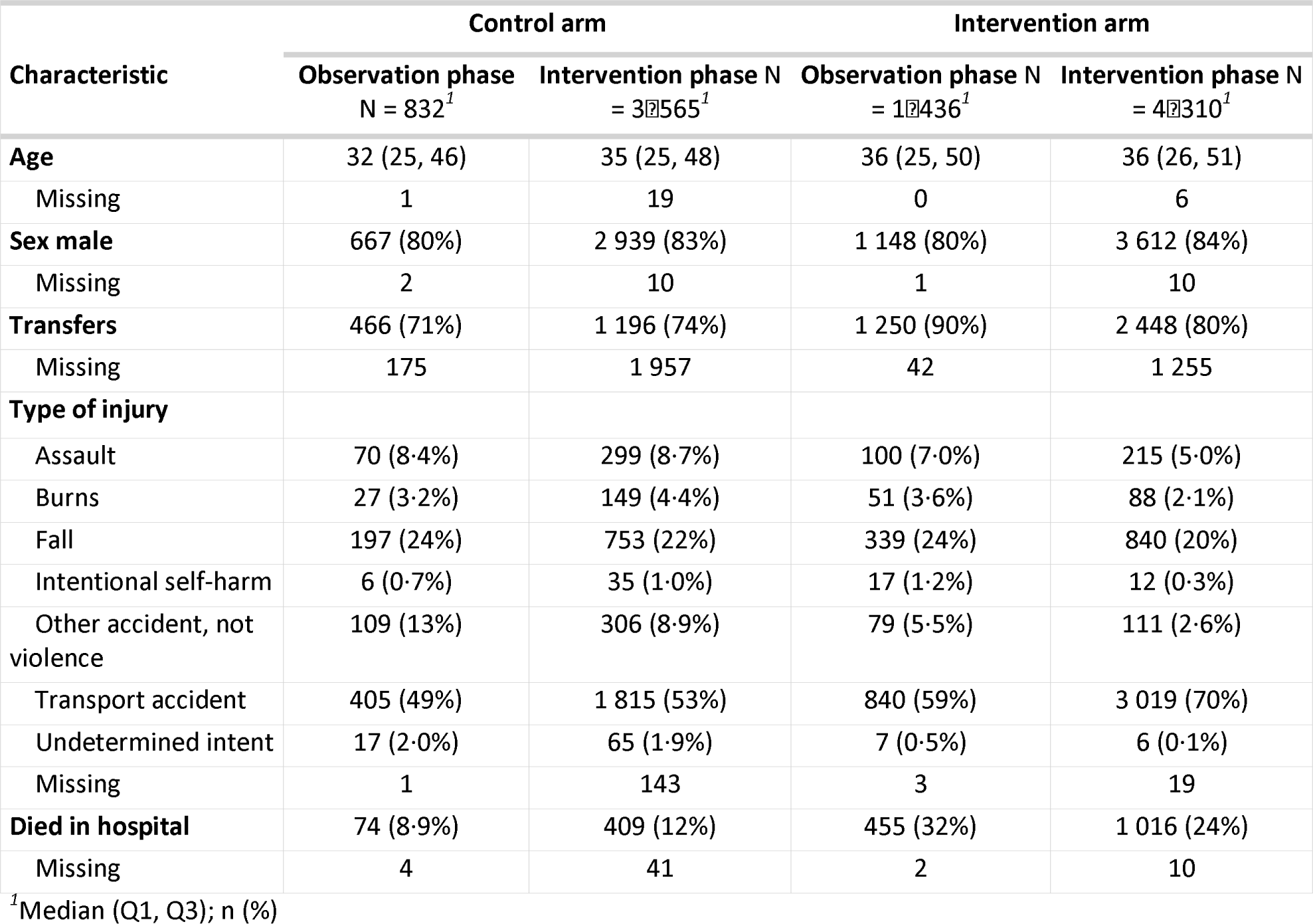
All included patients, n = 10143.

**Table 2:** All prospectively included patients, n = 4126.

| Characteristic | Control arm |  | Intervention arm |  |
| --- | --- | --- | --- | --- |
|  | Observation phase<br>N = 583<sup>1</sup> | Intervention phase<br>N = 888<sup>1</sup> | Observation phase<br>N = 938<sup>1</sup> | Intervention phase N<br>= 1717<sup>1</sup> |
| **Age** | 31 (24, 45) | 34 (25, 46) | 40 (25, 55) | 37 (27, 52) |
| Missing | 1 | 2 |  |  |
| **Sex male** | 470 (81%) | 720 (81%) | 729 (78%) | 1 438 (84%) |
| **Glasgow coma scale** | 15 (15, 15) | 15 (15, 15) | 15 (8, 15) | 12 (9, 15) |
| Missing | 17 | 38 | 5 | 6 |
| **Shock** | 5 (0.9%) | 18 (2.1%) | 54 (5.9%) | 25 (1.5%) |
| Missing | 22 | 34 | 20 | 14 |
| **Injury Severity Score (ISS)** | 6 (8) | 7 (8) | 12 (8) | 13 (7) |
| Missing | 50 | 134 | 164 | 166 |
| **Major trauma** | 103 (19%) | 155 (21%) | 486 (65%) | 1 164 (75%) |
| Missing | 53 | 142 | 186 | 174 |
| **CT done** |  |  |  |  |
| Before arrival | 11 (2.4%) | 32 (4.6%) | 506 (54%) | 833 (51%) |
| No | 219 (48%) | 302 (43%) | 137 (15%) | 101 (6.2%) |
| Yes | 227 (50%) | 363 (52%) | 293 (31%) | 688 (42%) |
| Missing | 126 | 191 | 2 | 95 |
| **Ultrasound done** |  |  |  |  |
| Before arrival | 0 (0%) | 1 (0.2%) | 2 (0.2%) | 31 (1.9%) |
| No | 258 (58%) | 341 (52%) | 908 (97%) | 1 455 (89%) |
| Yes | 187 (42%) | 312 (48%) | 22 (2.4%) | 147 (9.0%) |
| Missing | 138 | 234 | 6 | 84 |
| **Intubated** |  |  |  |  |
| Before arrival | 8 (2.0%) | 6 (1.1%) | 2 (0.2%) | 2 (0.1%) |
| No | 350 (89%) | 496 (90%) | 907 (97%) | 1 203 (76%) |
| Yes | 34 (8.7%) | 50 (9.1%) | 24 (2.6%) | 377 (24%) |
| Missing | 191 | 336 | 5 | 135 |
| **Admitted to ICU** | 62 (16%) | 109 (20%) | 13 (1.4%) | 77 (4.9%) |
| Missing | 197 | 340 | 12 | 148 |
| **Dead at 24 hours after arrival** | 14 (2.4%) | 17 (1.9%) | 99 (11%) | 109 (6.3%) |
| Missing | 4 | 7 | 1 | 0 |
| **Died in hospital** | 42 (7.3%) | 79 (9.1%) | 300 (32%) | 405 (24%) |
| Missing | 4 | 18 | 1 | 0 |
| **Dead at 30 days after arrival** | 46 (8.5%) | 85 (11%) | 328 (39%) | 435 (26%) |
| Missing | 43 | 94 | 102 | 20 |

<sup>1</sup>Median (Q1, Q3); n (%); Mean (SD)

In the intervention arm, we prospectively screened and collected structured data on 1454 additional patients for audit filter violations to inform discussions in the multidisciplinary review meetings. A list of audit filters with the most violations is available in the supplementary material. Descriptively, in the intervention arm, intubations rose from 2.6% to 24%, ultrasound increased from 2.4% to 9% and computed tomography (CT) scanning from 31% to 42% indicating changes in clinical practice aligned with key audit filters following implementation of the trauma quality improvement programme (Table 1).

### Time series analysis for in-hospital mortality Primary analysis

In the interrupted time series analysis, including 10,086 patients, the intervention phase was associated with a significant reduction in the odds of in-hospital mortality in the intervention arm *(OR: 0.56, 95% CI 0.4 to 0.77, p<0.001)*, corresponding to an estimated absolute difference of −11.2% *(95% CI −16% to −5.5%, p<0.001)*. No significant change was observed in the control arm *(OR: 0.94, 95% CI 0.5 to 1.75, p=0.83)*, absolute difference −0.5% *(95% CI −4% to 5.4%, p=0.835)*(Figure 2).

**Figure 2:**
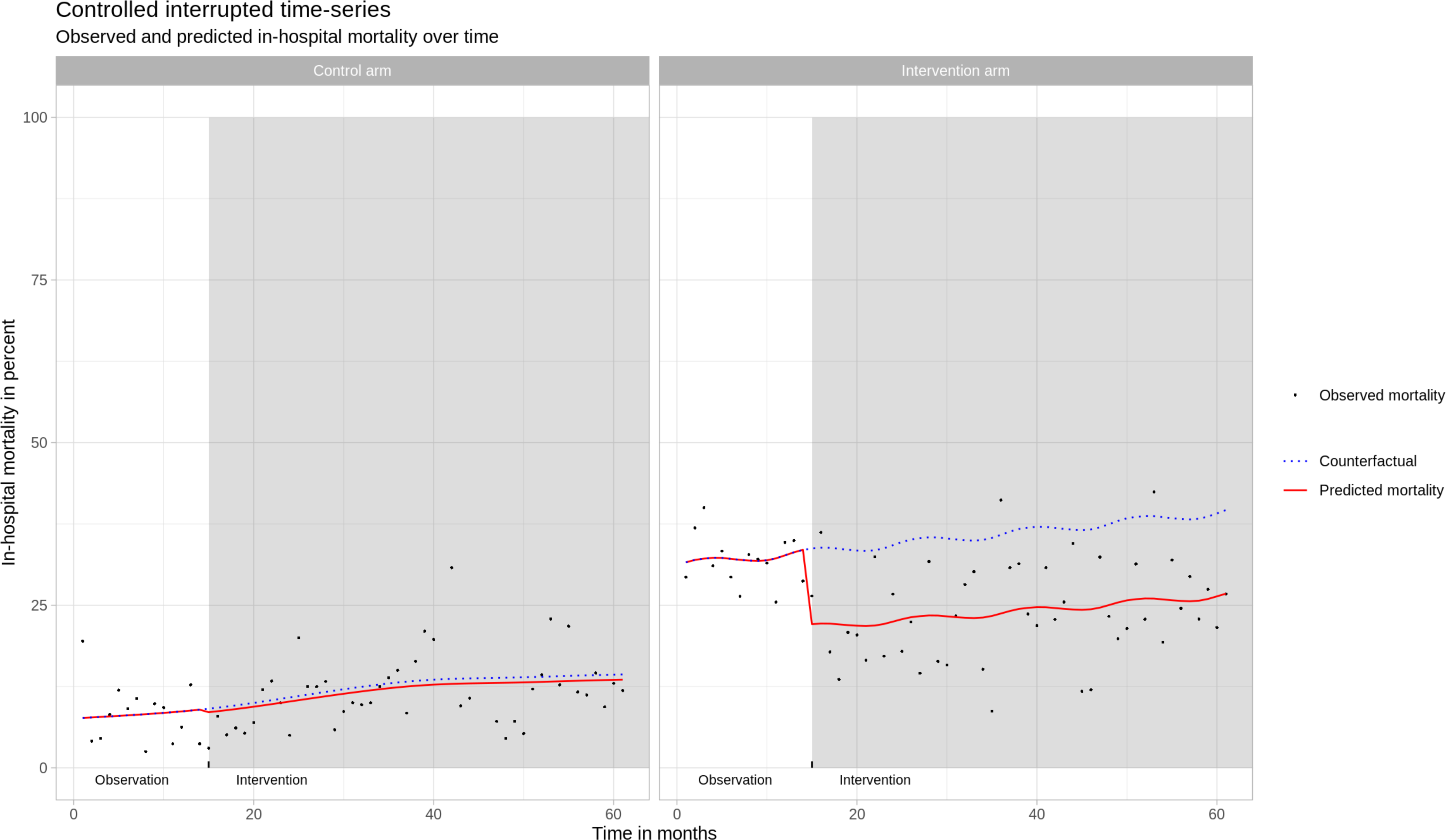
Interrupted time-series analysis.

The study time variable was not statistically significant *(p=0.14 for intervention; p=0.14 for control)*, suggesting that mortality remained otherwise stable over time, with no indication of other time-dependent trends influencing outcomes. We found no significant seasonal effects *(p=0.26 for intervention; p=0.48 for control)* and no evidence of autocorrelation in either study group *(p = 0.51 for intervention; p = 0.10 for control)*.

### Secondary and subgroup analysis

Formally comparing the control and intervention arms in the difference-in-differences analysis, including 3317 prospectively included patients, we found that the intervention was associated with a −12% difference in the in-hospital mortality rate *(−0.12, 95% CI - 0.16 to −0.09, p<0.001)* and a −15% difference in the 30-day mortality rate *(−0.15, 95% CI −0.19 to −0.11, p<0.001)* in the intervention arm compared to the control arm after adjusting for sex, age, GCS score, Injury Severity Score (ISS) and shock. For major trauma and potentially salvageable trauma patients, the reduction in mortality was more pronounced. No significant mortality change was observed in the control arm for either group (Table 3).

**Table 3:** Difference-in-differences analysis n=3317.

| Characteristic | All patients(n=3317) |  |  | Major trauma patients(n=1714) |  |  |  | Potentially salvageable trauma patients(n=1505) |  |  |  |
| --- | --- | --- | --- | --- | --- | --- | --- | --- | --- | --- | --- |
|  | Beta | 95% CI | p-value | Beta | 95% CI | p-value | Adj. p-value | Beta | 95% CI | p-value | Adj. p-value |
| Outcome: In-Hospital Mortality |  |  |  |  |  |  |  |  |  |  |  |
| Intervention Effect | -0.129 | -0.186 to -0.072 | <0.001 | -0.360 | -0.505 to -0.215 | <0.001 | <0.001 | -0.312 | -0.481 to -0.144 | <0.001 | <0.001 |
| Intervention Effect | -0.124 | -0.163 to -0.085 | <0.001 | -0.239 | -0.330 to -0.147 | <0.001 | <0.001 | -0.249 | -0.357 to -0.140 | <0.001 | <0.001 |
| Sex | -0.003 | -0.026 to 0.020 | 0.8 | -0.019 | -0.056 to 0.017 | 0.3 |  | 0.002 | -0.035 to 0.039 | >0.9 |  |
| Age | 0.001 | 0.001 to 0.002 | <0.001 | 0.002 | 0.001 to 0.002 | <0.001 |  | 0.002 | 0.001 to 0.002 | <0.001 |  |
| Injury Severity Score (ISS) | 0.001 | 0.000 to 0.002 | 0.2 | -0.007 | -0.010 to -0.004 | <0.001 |  | 0.007 | -0.008 to 0.022 | 0.4 |  |
| Glasgow coma scale | -0.083 | -0.086 to -0.080 | <0.001 | -0.092 | -0.096 to -0.088 | <0.001 |  | -0.090 | -0.094 to -0.086 | <0.001 |  |
| Shock | 0.072 | 0.012 to 0.132 | 0.018 | 0.020 | -0.055 to 0.096 | 0.6 |  | 0.084 | -0.002 to 0.169 | 0.055 |  |
| Outcome: 30 Day Mortality |  |  |  |  |  |  |  |  |  |  |  |
| Intervention Effect | -0.155 | -0.213 to -0.096 | <0.001 | -0.362 | -0.508 to -0.215 | <0.001 | <0.001 | -0.324 | -0.496 to -0.151 | <0.001 | <0.001 |
| Intervention Effect | -0.149 | -0.189 to -0.110 | <0.001 | -0.239 | -0.332 to -0.146 | <0.001 | <0.001 | -0.259 | -0.368 to -0.149 | <0.001 | <0.001 |
| Sex | 0.000 | -0.023 to 0.024 | >0.9 | -0.013 | -0.050 to 0.025 | 0.5 |  | 0.002 | -0.036 to 0.039 | >0.9 |  |
| Age | 0.001 | 0.001 to 0.002 | <0.001 | 0.002 | 0.001 to 0.002 | <0.001 |  | 0.002 | 0.001 to 0.002 | <0.001 |  |
| Injury Severity Score (ISS) | 0.001 | 0.000 to 0.002 | 0.13 | -0.006 | -0.009 to -0.003 | <0.001 |  | 0.005 | -0.010 to 0.021 | 0.5 |  |
| Glasgow coma scale | -0.085 | -0.088 to -0.082 | <0.001 | -0.093 | -0.097 to -0.089 | <0.001 |  | -0.093 | -0.097 to -0.089 | <0.001 |  |
| Shock | 0.075 | 0.014 to 0.136 | 0.017 | 0.017 | -0.060 to 0.093 | 0.7 |  | 0.082 | -0.004 to 0.168 | 0.063 |  |

Abbreviation: CI = Confidence Interval

### Sensitivity analysis

In the unadjusted pre-post analysis of all patients, the intervention was associated with an 8.0% absolute reduction in in-hospital mortality *(95% CI −11% to −5.3%, p<0.001)* and in the prospectively included cohort a 8.5% reduction in in-hospital mortality *(95% CI −12% to −4.8%, p<0.001)* and a 14% reduction in 30-day mortality *(95% CI −18% to - 9.5%, p<0.001)*. No statistically significant change was observed in the control arm.

Excluding the implementation phase and the COVID-19 period showed results consistent with the main analysis, with a statistically significant reduction in in-hospital mortality in the intervention arm. Excluding the trauma centre period resulted in a non-significant reduction; however, this analysis was limited to nine months of post-intervention data, including the six-month implementation phase. The adjusted analysis in the prospectively included intervention cohort showed a statistically significant reduction in both in-hospital and 30-day mortality after adjusting for age, sex, ISS, GCS, and shock. Full results are available in the supplementary material.

## Discussion

Our results indicate that implementing a trauma quality improvement programme using audit filters was associated with a substantial, significant reduction in-hospital mortality of 11%. We also observed increased use of ultrasound, intubations and CT scans, reflecting changes in care processes. However, the largest hospital in the intervention arm opened a trauma centre equipped with in-house CT scanner, multiple operating rooms, improved resuscitation capabilities and an increased number of ICU beds nine months after the beginning of the implementation phase. Although our sensitivity analysis revealed a statistical non-significant decrease in mortality before the trauma centre opened, it remains challenging to separate the impacts of the trauma centre and the quality improvement programme or determine whether their effects were synergistic. Still, our study shows that trauma mortality in this setting can be significantly reduced through system-level interventions.

Health systems are increasingly being recognised as complex adaptive systems, composed of interdependent agents and processes whose interactions give rise to emergent and often unpredictable outcomes.^24,25^ In such systems, effects are non-linear, and identifying direct causal chains between intervention and outcome is inherently difficult. We found that mortality declined after the programme was introduced, but the precise mechanisms responsible for this effect are challenging to disentangle. Although we did not formulate an explicit programme theory in advance, we believe that enabling health care staff to follow the trajectory of individual patients and their outcomes, and to review the processes of care, helped them to identify opportunities for improvement in their own context.

Several studies have reported improved mortality, particularly in severely injured patients, after trauma quality improvement implementation.^8,26–29^ However, definitions of trauma quality improvement vary, with heterogeneous interventions reported. Two studies conducted over 20 years ago in Germany and Thailand used trauma audit filters with a similar approach as our study, and reported reduced mortality and improved care processes.^30,31^ All were cohort studies using pre-post design. Development of audit filters for trauma has been done in more recent years, one study from Ghana and one from Cameroon.^32,33^. These filters differ substantially from those initially developed by ASCOT and later suggested in the WHO guidelines. In our previous study, the usefulness of the filters developed in Ghana and Cameroon was deemed high in India, compared filters developed in HICs.^16^

In HIC trauma quality improvement programmes and audit filters have been criticized for being inefficient in detecting opportunities for improvement and being costly.^34^ In obstetric care, audit processes have shown to improve outcomes while also emphasizing the importance of adaptation^35^, highlighting that standards of care need to be developed considering local priorities and knowledge to ensure that improvement efforts are directed to areas in greatest need. This is particularity important in settings with less developed trauma systems.

The maturity level of a trauma system is linked to reduced mortality and preventable deaths.^36,37^ Therefore, the effectiveness of these programmes in reducing mortality likely varies with system maturity. In a similar urban Indian setting, over 50% of trauma deaths were estimated to be preventable.^38^ Our results can likely be generalized to settings with similarly mature trauma systems, though local adaptations and contextual factors greatly influence their success. The core process of data collection and multidisciplinary case review can be utilized to empower providers to identify areas for improvement, learning, and education to develop the care provided. This can be conducted for trauma systems at all levels, even though data quality is a known barrier to this in LMICs.^8,39,40^

It should also be recognised that while this intervention, which is a scaffold for data collection and peer review, is introduced into a complex system. Regognizing health care as a complex adaptive system is important as effects are non-linear, What we can show ere is that that when this scafflod was introduced, mortality did appear to decline. Exactly by which mechanisms or the causla relationship between the intervention and the outcome is per definition in a complex system, challenging to identify. We did not have an explicit pre-porgram theory, we do belive that the ability for health care staff to understand the patient outcomes, follow the process of care for individual patients, helped them understand how to imporve care in the context where they work.

Our study has several limitations. First, the introduction of a new trauma centre at the largest intervention hospital may have influenced outcomes directly or acted synergistically with the trauma quality improvement programme. This hospital also accounted for a larger proportion of patients in the intervention arm, thus contributing more to the aggregate outcome estimates than the smaller intervention hospital.

Second, there was an unexpected difference in baseline mortality between the study arms. However, in the primary analysis, each arm was modeled independently, and the control arm was included to account for background trends over time and external events, rather than for direct comparison. In the secondary analysis, we used a difference-in-differences model, which assumes parallel pre-intervention trends rather than equivalent baseline levels. This allowed us to compare outcome changes over time between arms despite baseline differences in mortality. Third, 45 of 61 months fell below the target inclusion for the primary analysis; however, all months contributed data, and the number of time points before and after the intervention exceeded recommended thresholds for interrupted time series analysis. While smaller monthly samples increased uncertainty and variability in monthly estimates, the observed mortality reduction remained robust across analyses.

Fourth, the COVID-19 pandemic significantly impacted the participating centres. The smaller intervention centre was converted to a COVID-19 hospital, reducing trauma patient intake. Quarantine restrictions also decreased the overall number of trauma patients. Despite extending the study by 18 months and adjusting for the pandemic’s effects in the sensitivity analysis, its long-term impacts were likely beyond our ability to fully account for. The pandemic led to the suspension of review meetings, necessitating a restart post-pandemic. Last, both intervention hospitals receive a high number of transferred patients at varying intervals after the time of injury, potentially introducing bias, as patients may have died before arriving at the study hospital.

The intervention was designed to provide a structure containing training in trauma quality improvement processes, data collection and multidisciplinary review boards. The ongoing review process and implementation of changes were led entirely by the local teams, without involvement from the core research team. Finally, although we tracked the number of cases reviewed and meetings conducted, and held weekly calls with all project officers to support operations, ensure data collection, and facilitate the generation of reports for review meetings, we did not apply a formal framework to evaluate implementation fidelity. This limits our insight into how consistently the intervention was sustained and engaged with over time.

Despite these limitations, this is the first quasi-experimental study attempting to assess the impact of quality improvement programmes using audit filters on patient outcomes. We conducted a broad and comprehensive statistical analysis that supports our main findings, which are also in alignment with previous research. We believe that there is knowledge on how to select and adapt audit filters and implement a review process, but our understanding of how this translates into changes in care and outcomes is lacking. We also need better understanding of the implementation and sustainability of these programmes in complex health care systems, including how to best tailor them to local needs and detect potential negative effects, especially for other patient populations also competing for care resources.

In conclusion, our results suggest that system-level changes and data-driven quality improvement programmes using audit filters may reduce mortality in trauma patients. However, the challenge to understand what makes these programmes effective, useful and sustainable in terms of improving outcomes remains.

## Data Availability

All data in the present study consists of sensitive patient information. To protect the privacy of the participants, this data cannot be released outside the scope of this research collaboration in its current form. We are in the process of creating an anonymised version of this data, which we aim to release openly. This anonymized dataset will be published alongside a data descriptor paper to facilitate open-access use.

## Acknowledgement

We would like to thank Johan von Schreeb, professor at the department of Global Public Health at the Karolinska Institutet and Charles Mock, Professor Emeritus of Surgery and Epidemiology, University of Washington for their valuable input during this project. A warm thank you to Manjula Ranagatti and all project officers that have dedicated their time and expertise to this project and ensuring the quality of data collection.

JB and MGW had full access to all the data in the study and takes responsibility for the integrity of the data and the accuracy of the data analysis.

## Author contributions

MGW conceived the study and has been the PI. MGW, KS, MK, NR, SD, LFT, and MP contributed to the design of the study. SD was the project manager during study execution. MGW, KS, MK, NR, SD, JB, MJ, SR, MLN, RS, AM, SC, GB, DB, TK contributed to the execution of the study. JB prepared the data and conducted the statistical analysis. MGW and MP reviewed the results of the statistical analysis. JB drafted the first version of the article. All authors contributed to interpretation of data and critical revisions of the work for important intellectual content. MGW is the guarantor.

## Funding

This work was supported by the Swedish Research Council (2016-02041).

## Competing interests

None declared.

## Notes

### Competing Interest Statement

The authors have declared no competing interest.

### Clinical Trial

NCT03235388

### Author Declarations

Informed consent to take part of the intervention was not deemed applicable, as it was a hospital level intervention. We were granted waivers of informed consent for recording vital signs, demographic parameters and in-hospital outcomes. We obtained written consent for telephonic follow-up. Ethical approvals were granted by the Swedish Ethical Review Authority, approved 2017-06-07 2017/930-31/2, as well as at all local ethical review boards for each participating hospital: Maulana Azad Medical College, MAMC - approved 2017-07-19 F.1/IEC/MAMC/(57/02/2017/No113. SSKM/IPGME&R, Kolkata - approved 2017-08-21, IPGME&R/IEC/2017/396. JJ Hospital, Mumbai - approved 2017-08-22, No.IEC/Pharm/CT/111/A/2017. St Johns, Bangalore - approved 2017-08-24, 160/2017.

### Summary of Updates

Revised Background and new information box about previous research.

