## Supplementary material for "Effect of a continuous trauma quality improvement programme on mortality in urban India: a non-randomised controlled trial"

Date: 02 June, 2025

#### All implemented audit filters

Table 1: All included audit filters at intervention sites 1 and 2

| **Characteristic** | **Implemented at site 1** N = 24[1] | **Implemented at site 2** N = 32[1] |
|---|---|---|
| Audit filter | | |
| Airway breathing and circulation assessed immediately on arrival of patient to the emergency department | 0 | 1 |
| Assessment of mouth/throat for foreign bodies and debris made in patient that has difficulty breathing, within 10 minutes of arrival to emergency department | 1 | 1 |
| AVPU for initial assessment and followed by sequential GCS | 0 | 1 |
| Basic airway manoeuvre assistance (i.e. jaw-thrust, oral or nasal airway, suction, removal of foreign object) performed for patient with difficulty or obstructed breathing. | 0 | 1 |
| Basic airway manoeuvre assistance (i.e. sweep, chin-lift-jaw-thrust, oral or nasal airway, suction) performed for patient with difficulty or obstructed breathing. | 1 | 0 |
| Blood components started within 4 hours of arrival to the emergency department if the patient has a Hb < 7g/dl. | 0 | 1 |
| Breathing assessment made within 15 min of arrival to emergency department | 1 | 0 |
| Breathing assessment made within 5 min of arrival to emergency department | 0 | 1 |
| Burn patient did receive 2-4 mL of crystalloid solution per kilogram body weight per percent body surface burn within 24 hours of injury | 1 | 0 |
| Burn patient did receive 4 mL of Ringer's lactate per kilogram body weight per percent body surface burn within 24 hours of injury | 0 | 1 |
| Chest tube placed within 30 min of patient arrival in patient with suspected or confirmed pneumo- or hemothorax and oxygen saturation less than 98% | 1 | 1 |
| Documentation of history and physical examination by a doctor | 1 | 1 |
| Examination for pneumo- hemothorax done by listening to both sides of the chest with a stethoscope within 5 minutes of patient arrival to emergency department | 1 | 1 |
| FAST exam performed within 30 minutes from arrival to the emegency department to exclude hemoperitoneum. | 1 | 1 |
| Head computerized tomography (CT) scan done within 2 hours of arrival at hospital for a non-transferred patient with Glasgow Coma Scale score < 8 and systolic blood pressure > 90 | 0 | 1 |
| Hourly GCS in the emergency department of trauma patients with a diagnosis of skull fracture, intracranial injury or spinal cord injury | 1 | 0 |
| I.v antibiotics given within 1 hour of arrival to the emergency department in a patient with an open fracture | 1 | 1 |

| **Characteristic** | **Implemented at site 1** N = 24[1] | **Implemented at site 2** N = 32[1] |
|---|---|---|
| Immobilization and imaging performed in a patient with suspected spine injury, within 4 hours of arrival to the emergency department | 1 | 0 |
| Immobilization within 10 minutes and imaging performed within 4 hours of arrival to the emergency department in a patient with suspected spine injury | 0 | 1 |
| Intubation performed in patient with a GCS score of 8 or less within 10 minutes of arrival to emergency department. | 0 | 1 |
| Intubation performed in patient with a GCS score of 8 or less within 30 minutes of arrival to emergency department. | 1 | 0 |
| Laparotomy done within 1 hour of arrival to the emergency department in a patient with abdominal injuries and systolic blood pressure <90 after fluid resuscitation | 0 | 1 |
| Large bore IV was placed within 5 min of patient arrival to the emergency department in patients with tachycardia (heart rate > 110) or hypotension (systolic blood pressure < 90) | 0 | 1 |
| Large bore IV was placed within 5 minutes of patient arrival to the emergency department | 1 | 0 |
| MESS or WHO Trauma scale used in prognosis mangled upper extremity | 1 | 0 |
| Operation for irrigation and debridement within 12 hours from arrival to emergency department for an open fracture | 1 | 0 |
| Operation for irrigation and debridement within 6 hours from arrival to emergency department in a hemodynamically stable patient with an open fracture | 0 | 1 |
| Operation for sub or epidural hematoma within 3 hours of arrival to emergency department | 0 | 1 |
| Operative treatment of gunshot wound to the abdomen | 1 | 1 |
| Oxygen therapy with simple face mask initiated in a patient whose SpO2 is less than 92% within 5 minutes of initial assessment of the patient | 0 | 1 |
| Patient assessed for hypovolemia using clinical examination, USG, FAST, or DPL within 15 minutes of arrival to the emergency department when presenting with hypotension and tachycardia or suspected intra-abdominal bleeding, femoral shaft fracture, or pelvic fracture. | 0 | 1 |
| Patient assessed for hypovolemia when presenting with hypotension and tachycardia or suspected intra-abdominal bleeding, femoral shaft fracture, or pelvic fracture. | 1 | 0 |
| Pressure applied to external bleeding at patient arrival to the emergency department, and maintained until definitive control is performed | 1 | 1 |
| Reduction and/or splinting with analgesia made for a long bone fracture within 2 hours of admission or prior to transfer | 1 | 0 |
| Response time in initiating definite treatment from arrival to the emergency department, by specialist department, within 1 hour from arrival to the emergency department | 0 | 1 |
| Response time of respective department in attending the call | 0 | 1 |

| **Characteristic** | **Implemented at site 1** N = 24[1] | **Implemented at site 2** N = 32[1] |
|---|---|---|
| Sample sent for blood group and cross match in patients with significant bleeding (heart rate > 110 or systolic blood pressure < 90) within 15 minutes of arrival to the emergency department | 0 | 1 |
| Senior attending physician alerted when airway is compromised, usage jaw thrust, chin lift, ORA/NPA, or suction to open airway | 1 | 0 |
| Senior attending physician alerted within 5 minutes of patient arrival to the emergency department when airway is compromised, usage jaw thrust, chin lift, ORA/NPA, or suction to open airway | 0 | 1 |
| Senior medical officer made aware of patient with difficulty breathing, or shock present at triage (HR >100, OR SBP <110)* or oxygen saturation <95% within 5 minutes of initial assessment | 1 | 1 |
| Sequential (every 30 minutes) GCS of trauma patients with a diagnosis of skull fracture, intracranial injury or spinal cord injury | 0 | 1 |
| Serial assessment of vitals and GCS after admission | 1 | 0 |
| Splinting with analgesia made for a long bone fracture within 30 minutes of admission or prior to transfer | 0 | 1 |
| The clinician did assess airway patency by asking the patient a question and listening for a response | 1 | 1 |
| Vital signs recorded within 5 minutes of arrival to emergency department (must include breathing assessment, heart rate, blood pressure, oxygen saturation if available) | 1 | 0 |
| Vital signs recorded within 5 minutes of arrival to emergency department (must include breathing assessment, heart rate, blood pressure, oxygen saturation). | 0 | 1 |

[1]n

#### Audit filters with most violations

Table 2: Audit filters with more then 20 violations during the study period

| Audit filter | Number of violations |
|---|---|
| FAST exam performed within 30 minutes from arrival to the emegency department to exclude hemoperitoneum. | 811 |
| Examination for pneumo- hemothorax done by listening to both sides of the chest with a stethoscope within 5 minutes of patient arrival to emergency department | 704 |
| Breathing assessment made within 5 min of arrival to emergency department | 254 |
| Serial assessment of vitals and GCS every 2 hours for the first 12 hours after admission | 209 |
| Hourly GCS in the emergency department of trauma patients with a diagnosis of skull fracture, intracranial injury or spinal cord injury | 166 |
| Vital signs recorded within 5 minutes of arrival to emergency department (must include breathing assessment, heart rate, blood pressure, oxygen saturation if available) | 159 |
| Respiratory rate recorded within 10 minutes of arrival to emergency department | 100 |
| The clinician did assess airway patency by asking the patient a question and listening for a response | 64 |

| Audit filter | Number of violations |
|---|---|
| Serial assessment of vitals and GCS for the first 12 hours after admission | 58 |
| Large bore IV was placed within 5 minutes of patient arrival to the emergency department | 55 |
| Operation for irrigation and debridement within 12 hours from arrival to emergency department for an open fracture | 49 |
| Chest tube placed within 30 min of patient arrival in patient with suspected or confirmed pneumo- or hemothorax and oxygen saturation less than 98% | 45 |
| Documentation of history and physical examination by a doctor | 41 |
| Intubation performed in patient with a GCS score of 8 or less within 30 minutes of arrival to emergency department. | 39 |
| Immobilization performed within 10 minutes of arrival to the emergency department in a patient with suspected spine injury | 34 |
| Imaging performed within 4 hours of arrival to the emergency department in a patient with suspected spine injury | 27 |
| Airway secured in patient with a GCS score of 8 or less within 10 minutes of arrival to emergency department | 24 |
| MESS or WHO Trauma scale used in prognosis mangled extremity | 23 |
| Basic airway manoeuvre assistance (i.e. sweep, chin-lift-jaw-thrust, oral or nasal airway, suction) performed for patient with difficulty or obstructed breathing. | 22 |

### Primary analysis - full table

Table 3: Result of the interrupted times series analysis for in-hospital mortality, n = 10086

| | **Intervention arm** | | | **Control arm** | | |
|---|---|---|---|---|---|---|
| **Characteristic** | **exp(Beta)** | **95% CI** | **p-value** | **exp(Beta)** | **95% CI** | **p-value** |
| **Intervention** | 0·57 | 0·41 to 0·79 | 0·0006 | 0·94 | 0·50 to 1·76 | 0·84 |
| **s(study_month)** | | | 0·19 | | | 0·15 |
| **s(month)** | | | 0·27 | | | 0·45 |

Abbreviation: CI = Confidence Interval

### Sensitivity analysis

Table 4: Result of the interrupted times series analysis for in-hospital mortality, excluding implementation phase

| | **Intervention** (n = 55 ) | | | **Control** (n = 52 ) | | |
|---|---|---|---|---|---|---|
| **Characteristic** | **exp(Beta)** | **95% CI** | **p-value** | **exp(Beta)** | **95% CI** | **p-value** |
| **Intervention** | 0·56 | 0·40 to 0·79 | 0·0008 | 1·45 | 0·90 to 2·34 | 0·13 |
| **s(study_month)** | | | 0·17 | | | 0·56 |
| **s(month)** | | | 0·86 | | | 0·15 |

Abbreviation: CI = Confidence Interval

Table 5: Result of the interrupted times series analysis for in-hospital mortality, excluding time after trauma centre opened

| | **Intervention** (n = 23 ) | | | **Control** (n = 23 ) | | |
|---|---|---|---|---|---|---|
| **Characteristic** | **exp(Beta)** | **95% CI** | **p-value** | **exp(Beta)** | **95% CI** | **p-value** |
| **Intervention** | 0·75 | 0·47 to 1·21 | 0·24 | 0·74 | 0·29 to 1·91 | 0·54 |
| **s(study_month)** | | | 0·37 | | | 0·27 |
| **s(month)** | | | 0·27 | | | 0·60 |

Abbreviation: CI = Confidence Interval

Table 6: Result of the interrupted times series analysisfor in-hospital mortality, excluding Covid-19 phase

| | **Intervention** (n = 53 ) | | | **Control** (n = 50 ) | | |
|---|---|---|---|---|---|---|
| **Characteristic** | **exp(Beta)** | **95% CI** | **p-value** | **exp(Beta)** | **95% CI** | **p-value** |
| **Intervention** | 0·56 | 0·39 to 0·80 | 0·0016 | 0·88 | 0·51 to 1·52 | 0·65 |
| **s(study_month)** | | | 0·30 | | | 0·023 |
| **s(month)** | | | 0·49 | | | 0·080 |

Abbreviation: CI = Confidence Interval

Table 7: Results for logistic regression for in-hospital mortality, prospective cohort, n patients = 2201)

| | **In-hospital mortality (Unadjusted)** | | | **In-hospital mortality (Adjusted)** | | |
|---|---|---|---|---|---|---|
| **Characteristic** | **exp(Beta)** | **95% CI** | **p-value** | **exp(Beta)** | **95% CI** | **p-value** |
| post_intervention | 0·26 | 0·14 to 0·47 | <0·0001 | 0·45 | 0·27 to 0·74 | 0·0015 |
| s(study_month) | | | 0·0022 | | | 0·23 |
| s(month) | | | 0·23 | | | 0·059 |
| iss | | | | 0·99 | 0·92 to 1·06 | 0·78 |
| gcstot | | | | 0·65 | 0·56 to 0·75 | <0·0001 |
| age | | | | 1·02 | 0·97 to 1·07 | 0·45 |
| sex | | | | 0·53 | 0·12 to 2·36 | 0·40 |
| chock | | | | 15·1 | 2·79 to 81·9 | 0·0016 |

Abbreviation: CI = Confidence Interval

Table 8: Results for logistic regression for 30-day mortality, prospective cohort, n patients = 2201)

| | **30-day mortality (Unadjusted)** | | | **30-day mortality (Adjusted)** | | |
|---|---|---|---|---|---|---|
| **Characteristic** | **exp(Beta)** | **95% CI** | **p-value** | **exp(Beta)** | **95% CI** | **p-value** |
| post_intervention | 0·32 | 0·18 to 0·55 | <0·0001 | 0·50 | 0·31 to 0·82 | 0·0062 |
| s(study_month) | | | 0·033 | | | 0·86 |
| s(month) | | | 0·35 | | | 0·31 |
| iss | | | | 1·01 | 0·94 to 1·08 | 0·80 |

| | 30-day mortality (Unadjusted) | | | 30-day mortality (Adjusted) | | |
|---|---|---|---|---|---|---|
| **Characteristic** | **exp(Beta)** | **95% CI** | **p-value** | **exp(Beta)** | **95% CI** | **p-value** |
| gcstot | | | | 0·68 | 0·59 to 0·79 | <0·0001 |
| age | | | | 1·02 | 0·97 to 1·07 | 0·45 |
| sex | | | | 0·60 | 0·14 to 2·56 | 0·49 |
| chock | | | | 7·54 | 1·38 to 41·2 | 0·020 |

Abbreviation: CI = Confidence Interval

Table 9: Result of pre/post analysis for in-hospital mortality, all included patients

| | Control arm | | | | | Intervention arm | | | | |
|---|---|---|---|---|---|---|---|---|---|---|
| **Characteristic** | **Intervention phase** N = 3 565[1] | **Observation phase** N = 832[1] | **Difference**[2] | **95% CI**[2] | **p-value**[2] | **Intervention phase** N = 4 310[1] | **Observation phase** N = 1 436[1] | **Difference**[2] | **95% CI**[2] | **p-value**[2] |
| **Died in hospital** | 411 (12%) | 74 (8·9%) | 2·7% | 0·44% to 5·0% | 0·029 | 1 025 (24%) | 457 (32%) | -8·0% | -11% to -5·3% | <0·0001 |
| Missing | 41 | 4 | | | | 9 | 2 | | | |

[1]n (%)

[2]2-sample test for equality of proportions with continuity correction

Abbreviation: CI = Confidence Interval

Table 10: Result of pre/post analysis for in-hospital and 30 day mortality, all prospectively included patients

| | Control arm | | | | | Intervention arm | | | | |
|---|---|---|---|---|---|---|---|---|---|---|
| **Characteristic** | **Intervention phase** N = 888[1] | **Observation phase** N = 583[1] | **Difference**[2] | **95% CI**[2] | **p-value**[2] | **Intervention phase** N = 1 717[1] | **Observation phase** N = 938[1] | **Difference**[2] | **95% CI**[2] | **p-value**[2] |
| **Died in hospital** | 79 (9·1%) | 42 (7·3%) | 1·8% | -1·2% to 4·8% | 0·26 | 406 (24%) | 301 (32%) | -8·5% | -12% to -4·8% | <0·0001 |
| Missing | 18 | 4 | | | | 0 | 1 | | | |
| **Dead at 30 days after arrival** | 86 (11%) | 46 (8·5%) | 2·2% | -1·1% to 5·6% | 0·21 | 436 (26%) | 328 (39%) | -14% | -18% to -9·5% | <0·0001 |
| Missing | 88 | 42 | | | | 19 | 101 | | | |

[1]n (%)

[2]2-sample test for equality of proportions with continuity correction

| | Control arm | | | | | Intervention arm | | | | |
|---|---|---|---|---|---|---|---|---|---|---|
| **Characteristic** | **Intervention phase** N = 888[1] | **Observation phase** N = 583[1] | **Difference**[2] | **95% CI**[2] | **p-value**[2] | **Intervention phase** N = 1 717[1] | **Observation phase** N = 938[1] | **Difference**[2] | **95% CI**[2] | **p-value**[2] |

Abbreviation: CI = Confidence Interval
